# Indoor dust as a matrix for surveillance of COVID-19 outbreaks

**DOI:** 10.1101/2021.01.06.21249342

**Authors:** Nicole Renninger, Nick Nastasi, Ashleigh Bope, Samuel J. Cochran, Sarah R. Haines, Neeraja Balasubrahmaniam, Katelyn Stuart, Aaron Bivins, Kyle Bibby, Natalie M. Hull, Karen C. Dannemiller

## Abstract

Ongoing disease surveillance is a critical tool to mitigate viral outbreaks, especially during a pandemic. Environmental monitoring has significant promise even following widespread vaccination among high-risk populations. The goal of this work is to demonstrate molecular SARS-CoV-2 monitoring in bulk floor dust and related samples as a proof-of-concept of a non-invasive environmental surveillance methodology for COVID-19 and potentially other viral diseases. Surface swab, passive sampler, and bulk floor dust samples were collected from rooms of individuals infected with COVID-19, and SARS-CoV-2 was measured with quantitative reverse transcription polymerase chain reaction (RT-qPCR) and two digital PCR (dPCR) methods. Bulk dust samples had geometric mean concentration of 159 copies/mg-dust and ranged from non-detects to 23,049 copies/mg-dust detected using ddPCR. An average of 88% of bulk dust samples were positive for the virus among detection methods compared to 55% of surface swabs and fewer on the passive sampler (19% carpet, 29% polystyrene). In bulk dust, SARS-CoV-2 was detected in 76%, 93%, and 97% of samples measured by qPCR, chip-based dPCR, and droplet dPCR respectively. Detectable viral RNA in the bulk vacuum bags did not measurably decay over 4 weeks, despite the application of a disinfectant before room cleaning. Future monitoring efforts should further evaluate RNA persistence and heterogeneity in dust. This study did not measure virus viability in dust or potential transmission associated with dust. Overall, this work demonstrates that bulk floor dust is a potentially useful matrix for long-term monitoring of viral disease outbreaks in high-risk populations and buildings.

**Importance:** Environmental surveillance to assess pathogen presence within a community is proving to be a critical tool to protect public health, and it is especially relevant during the ongoing COVID-19 pandemic. Importantly, environmental surveillance tools also allow for the detection of asymptomatic disease carriers and for routine monitoring of a large number of people as has been shown for SARS-CoV-2 wastewater monitoring. However, additional monitoring techniques are needed to screen for outbreaks in high-risk settings such as congregate care facilities. Here, we demonstrate that SARS-CoV-2 can be detected in bulk floor dust collected from rooms housing infected individuals. This analysis suggests that dust may be a useful and efficient matrix for routine surveillance of viral disease outbreaks.

## Introduction

The spread of the novel severe acute respiratory syndrome coronavirus 2 (SARS-CoV-2) reached pandemic designation in March 2020 and has since resulted in more than 75 million cases of COVID-19 and 1.6 million deaths documented worldwide as of December 21, 2020 (1). Both symptomatic and asymptomatic carriers shed the virus into the environment (2–4). Viral particles are emitted primarily via respiratory droplets and aerosols, and persist on surfaces indoors (4–6). SARS-CoV-2 viability has been characterized after deposition onto several surface types (6). In one study, viable virus was detected on plastics and stainless steel up to 72 hours after application (5). Other studies have demonstrated respiratory viruses can contaminate environmental dust near infected individuals (7–9). These viral shedding routes together with persistence indoors and in environmental dust implicate potential viral contamination of indoor dust near infected individuals (10).

There is a critical need for targeted, efficient, and inexpensive methods to monitor SARS-CoV-2 and other viruses long-term to identify potential viral outbreaks prior to extensive spread. Fecal shedding of SARS-CoV-2 provides the basis for large-scale viral monitoring in wastewater systems (11–14). However, more targeted monitoring efforts are needed for indoor environments, especially those housing vulnerable populations such as congregate care facilities. We propose that the detection of SARS-CoV-2 RNA in indoor dust can be used for continued environmental surveillance of novel coronavirus, SARS-CoV-2. Targeted monitoring of dust in high-concern buildings could complement broader population-level monitoring approaches. This strategy could then be extended to other viruses of concern. Our goal is to demonstrate that indoor dust can be used as a matrix for viral outbreak surveillance.

## Results

We measured SARS-CoV-2 using quantitative reverse transcription polymerase chain reaction (RT-qPCR), chip-based digital PCR (dPCR), and droplet digital PCR (ddPCR) in samples of bulk dust, passive surface samples, and surface swabs from rooms of individuals with COVID-19. In bulk dust, the SARS-CoV-2 viral concentration had a geometric mean value of 159 copies/mg-dust and ranged from non-detects to 23,049 copies/mg-dust (Figure 1A). We detected SARS-CoV-2 RNA in 89% of bulk dust, 55% of surface swabs, and 38% of passive surface sampler samples (average among all three detection methods used). The ddPCR method detected viral RNA in 97% of bulk dust samples compared to 93% for the chip-based dPCR and 76% for RT-qPCR (Figure 1B).

**Figure 1:**
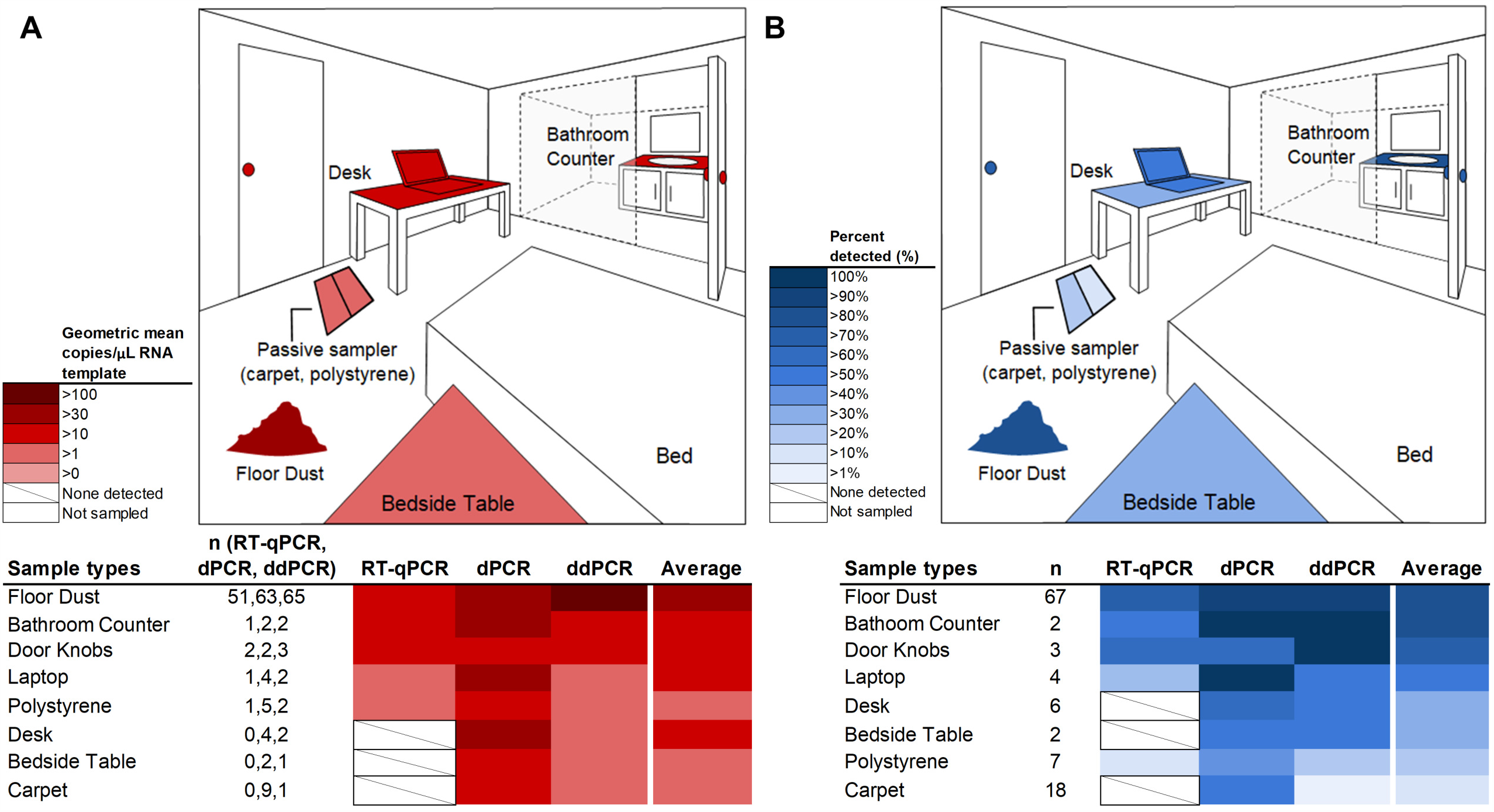
Heatmap displaying (A) the geometric mean copies/µL RNA template of samples above detection limit (samples below detection limit were excluded) and (B) the proportion of samples positive (%) for bulk dust, surface swabs, and passive sampler as observed with three PCR-based methods. The variable n is the number of samples used to calculate the geometric mean (positive detects) in (A) or proportion of samples positive in (B). Items in white on the room diagram were not sampled, and items in white with a slash on the lower heatmaps were not detected. Colors on the room diagrams represent the average value among all three measurement methods.

The COVID-19 isolation rooms were treated with a chlorine-based disinfectant prior to dust collection as part of the normal cleaning process, and the disinfectant is expected to largely inactivate the virus through reactions with the viral capsid (15). The bags were stored in the laboratory at room temperature after collection. Triplicate subsamples were extracted and viral RNA measured immediately upon collection and once per week for 4 weeks. Viral RNA did not measurably decay over 4 weeks in the vacuum bags (regression R^2^=0.009, p=0.47) (Figure 2A). The coefficient of variance (CoV) for copies/mg-dust ranged from 73.5-313.4% within each vacuum bag when averaged across the three methods of viral detection. This large variation in viral concentration is likely due to the heterogeneous mixture in the bags (Figure 2B-D).

**Figure 2:**
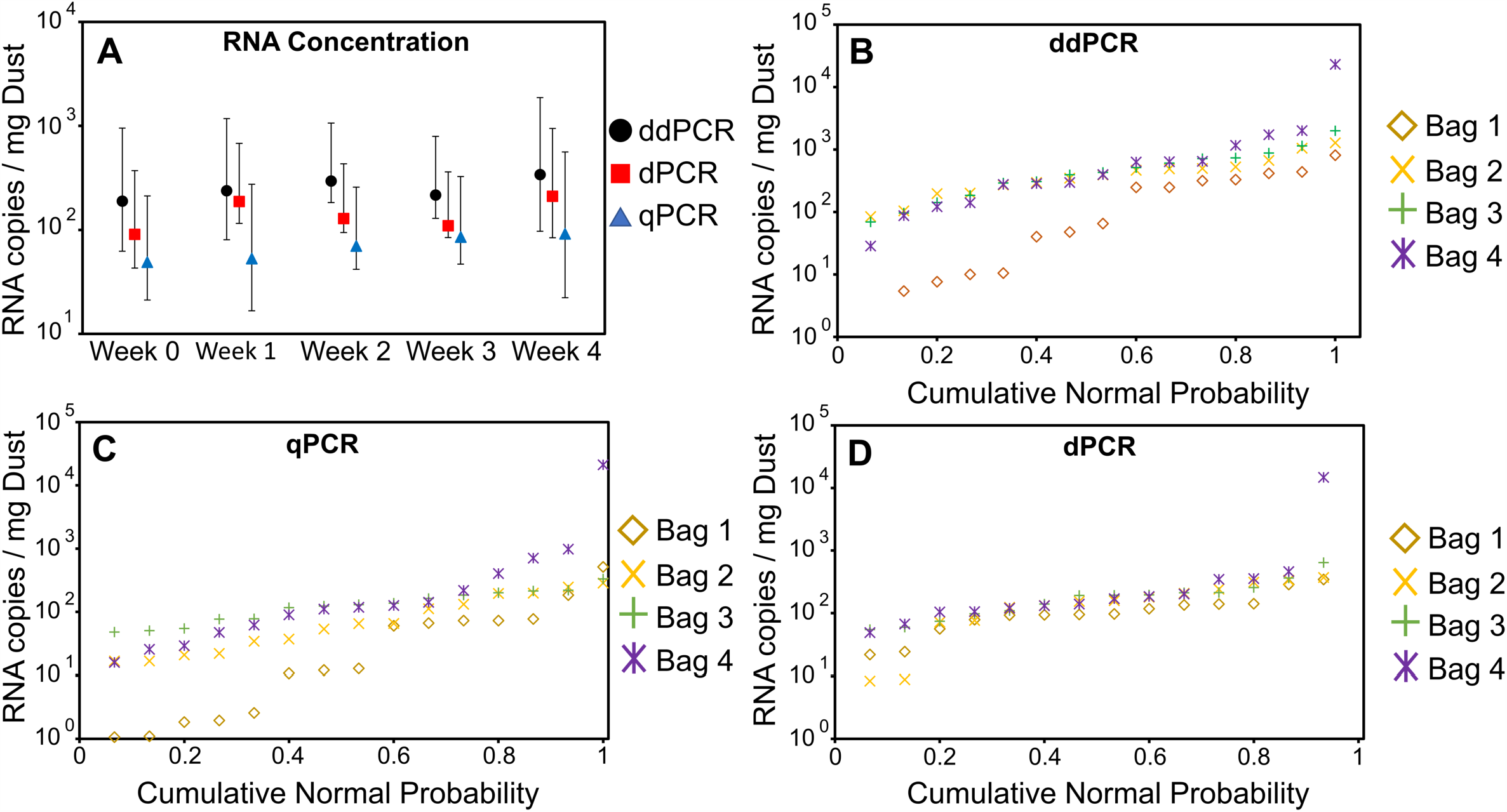
(A) RNA concentration of bulk dust samples (average of 4 bags) from initial collection to 4 weeks as measured by ddPCR, dPCR, and qPCR. Error bars shown represent the 95% confidence interval of each measurement. (B-D) Cumulative normal probability plots for each measurement method show variability of RNA concentration values for each bulk bag collected.

## Discussion

The novel coronavirus and the ongoing COVID-19 pandemic have highlighted the need for sensitive and scalable viral surveillance within communities. In the long term, the threat of COVID-19 outbreaks will subside to a level where indefinite routine testing of asymptomatic individuals may be too cumbersome or expensive. However, there will continue to be a need to more broadly monitor vulnerable populations such as those in long-term care facilities or high-risk patients in hospitals for SARS-CoV-2, influenza, respiratory syncytial virus (RSV), and other emerging viral diseases. Novel pathogens can be targeted with adaptable PCR-based assays. After detection, outbreaks can then be addressed with more targeted resources such as direct patient testing.

Our results demonstrate that environmental dust collection may provide a convenient and useful matrix for ongoing viral monitoring. The process can provide monitoring for many high-risk individuals, and dust samples are already being collected through normal cleaning practices such as vacuuming. Our observations indicate SARS-CoV-2 RNA in dust can persist at least four weeks after dust collection and the measured concentration can vary in different dust fractions within a vacuum bag. Therefore, multiple samples should be taken from a bag to more rigorously quantify the viral genetic signal or homogenization methods should be developed that comply with biosafety standards. Additionally, RNA and dust persistence in the environment should be considered when determining if the outbreak occurred recently or in the past.

Previously, measurements of indoor environmental microbes have been used to detect infectious microbes such as *Aspergillus fumigatus* and *Legionella pneumophila* (16–18). However, nucleotide-based tests do not measure infectivity, meaning the detection of genetic material from these microbes may indicate that people in the area are infected, but would not necessarily substantiate the risk of infection due to contact with indoor surfaces.

Indoor dust may also be used to complement other environmental surveillance methods, e.g. wastewater monitoring. Wastewater detection may be more beneficial at larger population scales covering thousands of individuals in a community, and one infected individual may be detectable among 100 to 2,000,000 individuals (19). Indoor dust may be useful in areas with smaller numbers of high-risk individuals where more specific outbreak identification is critical. Additionally, not all individuals secrete virus in stool (20). Indoor dust sampling may also be less expensive and be easier to implement, with simplified sample collection and no pre-concentration steps of samples required.

Limitations of this study include that we did not measure the viability of SARS-CoV-2 in the dust samples due to biosafety constraints, although this is not needed for surveillance. Also, our small sample size from rooms occupied by infected students may not be representative of other buildings and occupancy conditions, and samples were collected after a known infection as opposed to before. More information is needed on how representative each dust sample would be for a specific population and different occupancy levels. We were unable to sieve or otherwise homogenize the dust due to biosafety concerns, which likely resulted in variability within vacuum bags (Figure 2B-D). Additionally, decay of viral RNA in dust on a floor may differ from decay of viral RNA in dust treated with disinfectant and stored in a vacuum bag.

## Conclusions

Indoor dust provides an important matrix for environmental surveillance of viral disease outbreaks. Infected humans shed virus into their surrounding environment, which becomes integrated into the dust. In many cases, dust is already being collected during routine cleaning and can easily be submitted for analysis. Overall, dust may be a useful and efficient matrix to provide identification of viral disease outbreaks in high-risk settings, such as congregate care facilities. Future research can validate these results on a broader scale and in different building types to better inform use of this technique to mitigate viral transmission.

## Materials and Methods

### Overview

Samples were collected from two different homes, as well as isolation rooms used to quarantine individuals who tested positive for SARS-COV-2. Bulk dust was collected from both homes and from student isolation rooms. Surface swabs and a passive sampler collection were completed in one home. Viral RNA was measured using RT-qPCR, chip-based digital PCR, and droplet digital PCR.

### House surface swabs and passive sampler collection

Surface and passive samples were collected at the end of the 10 day quarantine period from two bedrooms of individuals who tested positive for SARS-CoV-2 in House 1. Surfaces were swabbed using sterile flocked swabs (Puritan, Maine, USA) and passive samplers consisting of carpet coupons and polystyrene coupons were placed on the floors of their isolation rooms. The passive samplers consisted of 3 cut pile carpet squares (fiber length 10 mm), 3 loop pile carpet squares (fiber length 7.5 mm), and 3 polystyrene squares attached to a template. All squares were 5 cm x 5 cm each. Both carpet types used were made of 100% polyethylene terephthalate (PET) fibers and a synthetic jute backing material. Fibers were specifically manufactured to contain no antimicrobial, stain, or soil resistance coatings. Swabs were dipped in autoclaved phosphate-buffered saline (PBS) and each was used on a different 10cm x 10cm surface area. Each wetted swab was wiped left and right across the 10cm x 10cm surface area, then rotated ⅓ turn and wiped to cover the surface area up and down, then rotated a final ⅓ turn and wiped in circular motions across the surface area. Swabs were placed back in the corresponding tube and resealed until extraction.

In Room 1, two swabs were used for the desk, two for a bedside table, and two for a computer. A passive sampler was placed on the floor by the bed for four days. In Room 2, two swabs were used for 100 cm2 areas on a computer, two for the same areas on a desk, two for the same areas on a second desk, one for the doorknob of the bedroom, two for 100 cm2 areas on the bathroom counter, and two on the bathroom doorknob, one for each side of the door. A passive sampler was placed on the floor between the desk and bed for two days, and another was placed on the open space in the bedroom for four days.

### House bulk dust

Bulk floor dust was collected from occupant vacuum bags of two different houses that had individuals infected with COVID-19. House 1 had floor dust collected 27 days after quarantine ended. House 2 had floor dust collected in the middle of the quarantine period.

### Isolation room bulk dust

Bulk dust samples were retrieved from four different vacuum bags used to clean the isolation rooms for students with COVID-19 at Ohio State University in Columbus, OH, USA and extracted over 4 weeks. Vacuum bags were collected by cleaning staff from rooms that were used to house students who tested positive for SARS-COV-2. One to two students would isolate in the rooms for 10 days after a positive diagnosis. The cleaning staff would vacuum and clean the rooms after the quarantine period was over and within 18 hours of the students leaving the rooms. Cleaning staff would spray the room with an electrostatic sprayer containing a disinfectant (sodium dichloro-s-triazinetrione, CAS 2893-78-9) and wait at least 20 minutes. This disinfectant provides free chlorine (stabilized by cyanuric acid), which non-selectively oxidizes biomolecules to inactivate pathogens. It is possible the disinfectant may be depleted by reacting with other organic material and biomolecules (dead skin, etc) and viral capsids in dust samples before impacting viral RNA (15). Cleaning staff used a Windsor Sensor XP12 vacuum (Kärcher, Denver, CO) to collect dust over a 3-4 week period. Each vacuum bag contained dust from approximately 30-50 isolation rooms as well as hallways, and potentially from surfaces in the isolation rooms if considered dusty. Isolation room flooring was vinyl composite tile and hallway flooring was wall to wall carpet.

### RNA Extraction

Viral RNA was extracted from dust and surface samples. Bulk dust samples and surface swabs were extracted using a Qiagen RNeasy Powermicrobiome extraction kit procedure (Qiagen, Hilden, Germany) modified to include 10x the recommended concentration of 2-mercaptoethanol and phenol based lysis. Triplicates of approximately 50 mg of dust were removed from bulk dust samples using an autoclaved spatula and each replicate extracted individually in a laminar flow biosafety cabinet. The spatula was flame sterilized between removing replicates. Bulk dust from student isolation rooms was stored in sealed bags and kept at a room temperature of approximately 22.8°C with a room relative humidity that fluctuated from 15-30%. Dust was not sieved due to biosafety concerns. Swabs were placed directly into the lysis tubes for extraction. Carpet samples from the passive sampler were extracted using the QIAmp DSP Viral RNA Mini Kit (Qiagen, Hilden, Germany). A 3cm x 1cm area was cut out of the middle of each carpet to reduce potential edge effects and vortexed for 1 minute in 4000 µL of autoclaved PBS. A total of 140 µL of this wash liquid was used in the RNA extraction. Swabs were dipped in autoclaved PBS and wiped horizontally and vertically across the polystyrene pieces on the passive sampler. All extraction sets included a blank to detect potential contamination. The RNA extract of a pre-pandemic dust sample collected in September 2019 was also tested and shown to be negative for SARS-CoV-2 on RT-qPCR with no amplification.

### Viral Detection

#### RT-qPCR

The viral detection assay targeted the N1 gene using the IDT SARS-CoV-2 (2019-nCoV) CDC qPCR Probe Assay (Integrated DNA Technologies, Inc., Coralville, IA, USA). This assay uses the 2019-nCoV_N1 forward primer (GAC CCC AAA ATC AGC GAA AT), the 2019-nCov_N1 reverse primer (TCT GGT TAC TGC CAG TTG AAT CTG) and the 2019-nCoV_N1 probe (FAM-ACC CCG CAT /ZEN/ TAC GTT TGG TGG ACC-3IABkFQ). Direct one-step real time qPCR amplification of cDNA was performed using qScript XLT One-Step RT-qPCR ToughMix (Quanta BioSciences, Gaithersburg, ML, USA). Each well contained 5µL of RNA template, 10 µL of qScript XLT One-Step RT-qPCR ToughMix, 1.5 µL of the IDT SARS-CoV-2 forward and reverse primers at 500 nM and probe at 125 nM, and 3.5 µL of sterile DI water. The 2019-nCoV plasmid control tenfold serial dilutions were used as a standard curve to calculate copies per µL of RNA template, based on plasmid quantification determined by dPCR (see below) (Integrated DNA Technologies, Inc., Coralville, IA, USA). Cycling parameters were set following the instructions supplied by the CDC for qScript XLT One-Step RT-qPCR ToughMix (21). Cycling parameters consisted of 10 min at 50°C for 1 cycle, 3 min at 95°C for 1 cycle, and 3 seconds at 95°C followed by 30 seconds at 55°C for 50 cycles. Seven no template controls were tested and no amplification occurred.

A subset of 10% of samples were tested for inhibition. RNA template was spiked with positive plasmid control to test for a reduction in signal due to the presence of inhibitors. The spike concentration was 100 times the highest sample concentration determined by qPCR. Inhibition was indicated if there was a delay in expected amplification. Each sample type was tested for inhibition: bulk dust, swab, and passive sampler. No inhibition was detected in any of the sample types except for the carpet wash from the passive sampler, where the inhibition delayed amplification by 1.45 cycles. Diluting these samples by 10 fold to reduce inhibition would place these samples below the detection limit of 2.3 copies per µL RNA.

#### Chip-based dPCR

Digital PCR was performed using the QuantStudio 3D Digital (QS3D) PCR System (Applied Biosystems, Forest City, CA) that utilizes a chip-based technology. This system uses a QuantStudio 3D Digital PCR Chip Adapter Kit for the ProFlex Flat Block Thermal Cycler equipped with a tilt base, which holds the chips (version 2) in place during thermocycling. cDNA was first reverse transcribed from RNA samples using the iScript cDNA Synthesis Kit (Biorad, Hercules, CA) according to the recommended reaction protocol on the ProFlex PCR System (Applied Biosystems, Forest City, CA). RNA was detected and quantified using the N1 assay described above. Each reaction was prepared as a 15 µL volume consisting of 2.00 µL water, 7.25 µL of QuantStudio 3D Digital PCR Master Mix v2 (Applied Biosystems, Forest City, CA), forward and reverse primers at 500 nM, probe at 125 nM, and 5 µL of RNA extract. 14.5 µL of the solution was transferred into the sample loading port of the loading blade and then loaded onto the chip. Immersion fluid was used to cover the surface of the chip. The chip was then sealed and additional immersion fluid was added to fill the chip case. Thermal cycling consisted of 10 minutes at 96°C, 39 cycles of 60°C for two minutes followed by 98°C for 30 seconds, and finally 60°C for 2 minutes. The cover temperature was set at 70°C and the reaction volume was set to 1 nL. Each experiment included one negative control and one N1 positive control (2019-nCoV_N_Positive Control, Integrated DNA Technologies, Inc., Coralville, IA, USA). Chips were then removed and imaged using the QuantStudio 3D Digital PCR Instrument following thermal cycling. Manual thresholding and quantification was performed using the QuantStudio 3D AnalysisSuit Software. The 95% limit of blank for the N1 assay was determined to be 1.45 gene copies per µL of reaction mixture using seven replicates of negative controls. Inhibition was not assessed for dPCR.

#### ddPCR

Droplet digital PCR was performed using the Bio-Rad QX200 system along with a C1000 Touch Thermal Cycler (Biorad, Hercules, CA). SARS-CoV-2 RNA was detected and quantified using the N1 assay previously described. Inhibition was assessed by spiking a subset of sample extracts (n=17) with bovine respiratory syncytial virus (BRSV) RNA extracted directly from a live attenuated bovine vaccine (Inforce 3 Cattle Vaccine, Zoetis, Parsippany-Troy Hills, NJ) using a Qiagen PowerViral AllPrep DNA/RNA kit (Hilden, Germany). BRSV RNA was detected and quantified using an assay targeting the nucleoprotein gene with forward primer (GCA ATG CTG CAG GAC TAG GTA TAA T), reverse primer (ACA CTG TAA TTG ATG ACC CCA TTC T), and probe (FAM-ACC AAG ACT/ZEN/TGT ATG ATG CTG CCA AAG CA-3IABkFQ) (Integrated DNA Technologies, Inc., Coralville, IA, USA) (22). Each N1 ddPCR reaction was prepared as a 22 µL volume consisting of 5.45 µL water, 5.45 µL of One-Step RT-ddPCR Supermix (Bio-Rad, Hercules, CA), 2.1 µL of Reverse Transcriptase, 1.05 µL Dithiothreitol, forward and reverse primers at 1000 nM, probe at 250 nM, and 5 µL of RNA extract. BRSV wells were prepared in the same manner except forward and reverse primers at 900 nM and probe at 250 nM. A volume of 20 µL of each reaction mixture was passed into droplet generation. Thermal cycling was performed with reverse transcription for 60 minutes at 50°C, followed by 10 minutes at 95°C, forty cycles of 95°C for 30 seconds followed by 59°C for 1 minute, and finally 98°C for 10 minutes. Each ddPCR experiment included two no-template controls each for BRSV and N1 and two positive controls each for BRSV (RNA and molecular water) and N1 (2019-nCoV_N_Positive Control, Integrated DNA Technologies, Inc., Coralville, IA, USA). Manual thresholding and quantification was performed in QuantaSoft Version 1.7.4 (Bio-Rad, Hercules, CA) such that no-template controls yielded no positive droplets.

The 95% limit of detection for the N1 assay was determined to be 3.3 gene copies per ddPCR reaction using a ten-replicate dilution series of synthetic SARS-CoV-2 RNA control material (MT188340, Twist Bioscience, San Francisco, CA) with a cumulative Gaussian distribution fit to the observed proportion of the replicates positive along the dilution gradient. There was no evidence of inhibition as no difference was observed in the quantification of BRSV RNA in sample extracts compared to the BRSV positive controls (two-tailed t-test, p=0.19).

### Statistical and data analysis

Our goal was to compare measurement of SARS-CoV-2 in bulk dust, on surface swabs, and on a passive sampler using three different measurement methods. Each vacuum bag of dust was sampled and extracted in triplicate at each time point (immediately after collection and 1,2,3 and 4 weeks post collection). All three detection methods (qPCR, dPCR and ddPCR) analyzed the same sample extractions for all sample types. Detection limit information is described above for each detection method. The geometric mean was reported for quantification of SARS-CoV-2 RNA present in samples using each method due to the logarithmic nature of PCR-based data. Potential RNA decay over the 4 week time period was evaluated in bulk dust with a regression analysis on the ddPCR data transformed with the natural logarithm. The dataset is available at https://doi.org/10.5061/dryad.3n5tb2rg1.

## Data Availability

Data will be available as indicated in the manuscript upon peer-reviewed publication.

https://doi.org/10.5061/dryad.3n5tb2rg1

## Acknowledgements

We are grateful to the isolation room coordinator/manager and staff, as well as the occupants for donation of the samples. We would also like to thank the carpet manufacturer for donation of well-characterized carpet samples for the passive sampler.

